# Shortwave infrared imaging increases the number of lymph nodes and the number of positive lymph nodes in surgical pathology specimens – a 104-patient study

**DOI:** 10.1101/2024.11.24.24317845

**Authors:** J M Jorns, R S Smith, D Strenk, G Stauble, S Khelifa, R Seltzer, J Manuel, M W Rosenbaum, J Almas, Z Li

## Abstract

**Aims:** Adequate lymph node examination is key to accurate cancer staging. Lymph node search in surgical resection specimens is a challenging task because it can be labor intensive and inconsistent. This study investigates the effectiveness of shortwave infrared imaging technology in increasing the overall and positive lymph node numbers.

**Methods:** 104 surgical resection specimens from two hospitals were first grossed with traditional manual palpation. The residual tissues were then grossed with the assistance of shortwave infrared imaging. The numbers of lymph nodes and positive lymph nodes within these two steps were documented and compared.

**Results:** In 90 of the 104 cases (86.5%), shortwave infrared imaging assisted gross examination identified additional lymph nodes. In 11 of 104 cases (10.6%), shortwave infrared imaging assisted gross examination identified additional positive lymph nodes.

**Conclusions:** Shortwave infrared imaging increases the number of lymph nodes and the number of positive lymph nodes in surgical pathology specimens compared to traditional visual and manual dissection.

## INTRODUCTION

Adequate lymph node staging is critically important for accurate cancer staging.(1) It has been widely shown that the number of lymph nodes examined is correlated with overall improved prognosis, possibly because of increased accuracy in staging. This association has been reported in colon cancer,(1) rectal cancer,(2) gastric cancer,(3) lung cancer,(4) head and neck cancers,(5) among others. As a result, different minimum numbers of lymph nodes have been proposed across many cancer types. The most well-known benchmark is a yield of at least 12 lymph nodes for colorectal cancer specimens.(6)

The task of finding lymph nodes is laborious, time-consuming, and produces inconsistent results.(7–9) This problem has even become a common point of friction between pathology and surgery departments.(10)

Attempts have been made to improve the efficiency and accuracy of lymph node dissection over the past few decades. The most notable solution is a method in which the specimen is soaked in organic solvents, such as acetone,(11) Carnoy’s solution,(12) and GEWF solution.(13) However, this procedure significantly extends turnaround time and requires a designated chemical handling protocol. As a result, it has not been adopted in many institutions for routine gross examination. Another alternative is to submit the specimens in entireties. This can be done for smaller specimens,(14) but is not feasible for large cases. Even for small cases, it is an inefficient and wasteful process that results in processing greater numbers of cassettes and increased microscopic review time.(14)

Shortwave infrared (1000 – 2000 nm) imaging is a novel emerging field for pathological imaging. The unique property of this wavelength range is that water, while being transparent in the visible light (400 – 700 nm) and near-infrared (700 – 1000 nm) range, is highly absorbent of shortwave infrared light. Therefore, when shortwave infrared light is illuminated onto biological tissues, the tissue types that are rich in water, such as lymph node and lymphatic vessels, will absorb the light and appear as dark areas. The tissue types that are low in water content, such as fat, will reflect the shortwave infrared light and appear as bright areas.

Cision InVision™ is an imaging device based on shortwave infrared imaging. Surgical specimens can be placed on the glass window of the imaging device and a reflection-based shortwave infrared image will be displayed on its monitor in real time. (Figure 1A) The prosector can manually palpate the surgical specimen and dissect out the lymph node candidates on the glass window. It is worth noting that the technique only displays a reflection image of shortwave infrared light highlighting the natural water content difference – it does not specifically highlight the lymph nodes for the prosectors. The prosector still needs to rely on one’s prior experience with lymph node grossing to determine whether the identified darker areas are lymph nodes, as opposed to vessels, or other structures that may display high relative water content. The purpose of this pilot study was to investigate the effectiveness of this novel imaging technology to improve lymph node yield and its impact on cancer staging.

**Figure 1.**
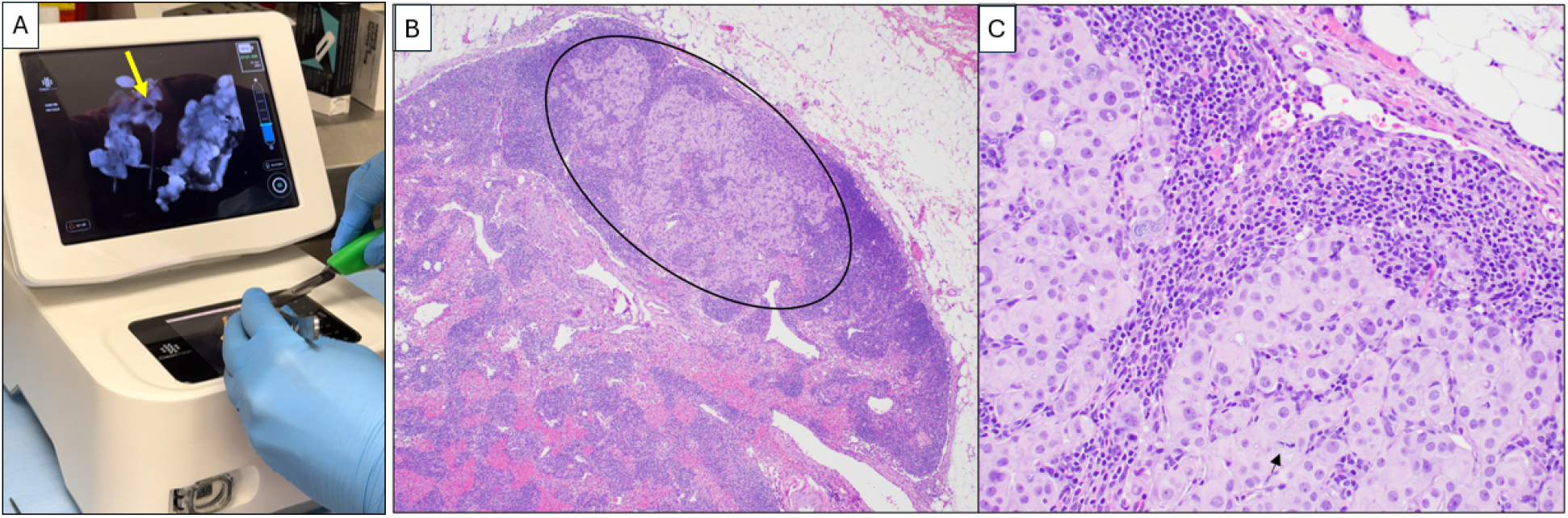
(A) A prosector using the Cision InVision™ shortwave infrared imaging device on a surgical resection specimen for lymph node dissection. (B) Example case (Case H1-2) from a breast cancer axillary lymph node dissection in which the only positive lymph node was identified via shortwave infrared imaging. Metastatic carcinoma is located within lymph node parenchyma (circle) (B: H&E, 4X), and consists of cohesive cells with large, pleomorphic nuclei, prominent nucleoli, and readily identifiable mitoses (arrow) (C: H&E, 20X).

## METHODS

### Patients

This is a pilot study where the effectiveness of shortwave infrared imaging for lymph node searches is evaluated for the first time. The study included 104 cancer patients from two hospitals, 37 cases from an academic hospital and 67 cases from a large community hospital. The cancer types for these cases include colon (68 cases), rectum (17 cases), breast (3 cases), pancreas (8 cases), stomach (5 cases), skin (1 case) and small bowel (2 cases).

### Pathologists and Pathologists’ Assistants

The gross examination procedures and the operation of the shortwave infrared imaging device were conducted by pathologists and board-certified Pathologists’ Assistants (PA) under the supervision of board-certified pathologists. Microscopic examinations of submitted lymph node candidates were confirmed by board certified pathologists.

### Study Procedure

To evaluate the effectiveness of shortwave infrared imaging in assisting lymph node searches compared to standard of care, the study is designed to have two steps:

In the first step, the gross examination for lymph nodes was performed with visual inspection and manual palpation. The lymph node candidates retrieved in this step were placed in the first batch of cassettes, Batch A. This step is representative of the current standard of care.

In the second step, the same PA, under the supervision of a pathologist, used the Cision InVision™ Pathology Imaging System to search for additional lymph nodes in residual specimens after the first step. The lymph node candidates that were retrieved in this step were placed in the second batch of cassettes, Batch B.

Batch A and B cassettes were processed via standard histological protocols. Microscopic review was performed by board-certified pathologists to confirm the total numbers of lymph nodes and the numbers of positive lymph nodes in Batches A and B.

### Statistics

Poisson regression was used to test the hypotheses that (1) the number of lymph nodes identified by shortwave infrared imaging was significantly different than zero, (2) the number of positive lymph nodes identified by shortwave infrared imaging was significantly different than zero.

This statistical technique was used to test hypotheses where the outcome scale was counted data. The two hypothesis outcomes comprised distinct counts of total lymph nodes detected, and distinct counts of positive lymph nodes detected. Statistical significance was concluded with Wald Chi Square p values less than 0.05. Analysis was run using the GENMOD procedure in SAS 9.4 (Cary, NC), with the Poisson distribution and the log-linear regression model specifications.

## Results

**Table 1.** Total lymph node and positive lymph node counts with traditional palpation and with assistance via shortwave infrared imaging in Hospital 1 (H1)

| Study Case Number | Disease Type | Total LN Traditional Palpation | Positive LN Traditional Palpation | Shortwave Assisted | Positive LN Shortwave Assisted | Improvement Percentage | Affect Staging | Affect Compliance |
| --- | --- | --- | --- | --- | --- | --- | --- | --- |
| H1-1 | Rectal | 8 | 1 | 13 | 1 | 162.50% | Y | Y |
| H1-2 | Breast | 6 | 0 | 5 | 1 | 83.33% | Y | N |
| H1-3 | Colon | 2 | 0 | 0 | 0 | 0.00% | N | N |
| H1-4 | Colon | 13 | 0 | 7 | 0 | 53.85% | N | N |
| H1-5 | Colon | 66 | 3 | 12 | 0 | 18.18% | N | N |
| H1-6 | Rectum | 1 | 0 | 6 | 0 | 600.00% | N | N |
| H1-7 | Colon | 28 | 0 | 1 | 0 | 3.57% | N | N |
| H1-8 | Colon | 13 | 0 | 5 | 0 | 38.46% | N | N |
| H1-9 | Rectum | 17 | 0 | 3 | 0 | 17.65% | N | N |
| H1-10 | Pancreas | 4 | 0 | 2 | 0 | 50.00% | N | N |
| H1-11 | Rectum | 3 | 0 | 4 | 0 | 133.33% | N | N |
| H1-12 | Rectum | 31 | 0 | 9 | 0 | 29.03% | N | N |
| H1-13 | Colon | 5 | 0 | 1 | 0 | 20.00% | N | N |
| H1-14 | Colon | 5 | 0 | 15 | 0 | 300.00% | N | Y |
| H1-15 | Colon | 5 | 0 | 2 | 0 | 40.00% | N | N |
| H1-16 | Rectum | 59 | 0 | 9 | 0 | 15.25% | N | N |
| H1-17 | Pancreas | 3 | 0 | 2 | 0 | 66.67% | N | N |
| H1-18 | Colon | 22 | 1 | 4 | 0 | 18.18% | N | N |
| H1-19 | Breast | 5 | 3 | 6 | 2 | 120.00% | Y | N |
| H1-20 | Skin | 23 | 0 | 2 | 0 | 8.70% | N | N |
| H1-21 | Colon | 8 | 0 | 12 | 0 | 150.00% | N | Y |
| H1-22 | Pancreas | 8 | 0 | 7 | 0 | 87.50% | N | Y |
| H1-23 | Stomach | 3 | 1 | 3 | 0 | 100.00% | N | N |
| H1-24 | Rectum | 5 | 0 | 4 | 0 | 80.00% | N | N |
| H1-25 | Colon | 24 | 0 | 2 | 0 | 8.33% | N | N |
| H1-26 | Pancreas | 21 | 0 | 4 | 0 | 19.05% | N | N |
| H1-27 | Colon | 4 | 0 | 28 | 0 | 700.00% | N | Y |
| H1-28 | Pancreas | 21 | 4 | 6 | 0 | 28.57% | N | N |
| H1-29 | Pancreas | 12 | 0 | 3 | 0 | 25.00% | N | N |
| H1-30 | Colon | 15 | 0 | 8 | 0 | 53.33% | N | N |
| H1-31 | Rectum | 25 | 0 | 11 | 0 | 44.00% | N | N |
| H1-32 | Rectum | 0 | 0 | 13 | 0 | N/A | N | Y |
| H1-33 | Pancreas | 13 | 0 | 5 | 0 | 38.46% | N | N |
| H1-34 | Colon | 8 | 0 | 17 | 0 | 212.50% | N | Y |
| H1-35 | Rectum | 7 | 0 | 12 | 0 | 171.43% | N | Y |
| H1-36 | Pancreas | 20 | 5 | 8 | 1 | 40.00% | Y | N |
| H1-37 | Colon | 32 | 0 | 12 | 0 | 37.50% | N | N |

**Table 2.** Total lymph node and positive lymph node counts with traditional palpation and with assistance via shortwave infrared imaging in Hospital 2 (H2)

| Study Case Number | Disease Type | Total LN Traditional Palpation | Positive LN Traditional Palpation | Shortwave Assisted | Positive LN Shortwave Assisted | Improvement Percentage | Affect Staging | Affect Compliance |
| --- | --- | --- | --- | --- | --- | --- | --- | --- |
| H2-1 | Colon | 10 | 0 | 11 | 0 | 110.00% | N | Y |
| H2-2 | Stomach | 11 | 1 | 5 | 1 | 45.45% | Y | Y |
| H2-3 | Colon | 8 | 0 | 7 | 0 | 87.50% | N | Y |
| H2-4 | Colon | 10 | 0 | 0 | 0 | 0.00% | N | N |
| H2-5 | Colon | 8 | 0 | 13 | 0 | 162.50% | N | Y |
| H2-6 | Colon | 18 | 8 | 10 | 1 | 55.56% | Y | N |
| H2-7 | Colon | 10 | 0 | 5 | 0 | 50.00% | N | Y |
| H2-8 | Stomach | 8 | 4 | 4 | 2 | 50.00% | Y | Y |
| H2-9 | Small Bowel | 0 | 0 | 5 | 1 | 100.00% | Y | N |
| H2-10 | Colon | 2 | 0 | 4 | 0 | 200.00% | N | N |
| H2-11 | Colon | 11 | 0 | 6 | 0 | 54.55% | N | Y |
| H2-12 | Colon | 6 | 0 | 4 | 0 | 66.67% | N | N |
| H2-13 | Colon | 5 | 0 | 4 | 0 | 80.00% | N | N |
| H2-14 | Colon | 6 | 0 | 1 | 0 | 16.67% | N | N |
| H2-15 | Colon | 13 | 4 | 3 | 0 | 23.08% | N | N |
| H2-16 | Colon | 10 | 0 | 0 | 0 | 0.00% | N | N |
| H2-17 | Colon | 2 | 0 | 8 | 0 | 400.00% | N | N |
| H2-18 | Colon | 19 | 0 | 4 | 0 | 21.05% | N | N |
| H2-19 | Colon | 6 | 0 | 0 | 0 | 0.00% | N | N |
| H2-20 | Colon | 10 | 2 | 1 | 0 | 10.00% | N | N |
| H2-21 | Colon | 7 | 1 | 0 | 0 | 0.00% | N | N |
| H2-22 | Colon | 4 | 1 | 0 | 0 | 0.00% | N | N |
| H2-23 | Breast | 1 | 0 | 5 | 1 | 500.00% | Y | N |
| H2-24 | Colon | 15 | 0 | 1 | 0 | 6.66% | N | N |
| H2-25 | Colon | 36 | 20 | 3 | 0 | 8.33% | N | N |
| H2-26 | Colon | 12 | 7 | 0 | 0 | 0.00% | N | N |
| H2-27 | Small Bowel | 0 | 0 | 4 | 0 | 100.00% | N | N |
| H2-28 | Colon | 6 | 0 | 5 | 0 | 83.33% | N | N |
| H2-29 | Rectum | 20 | 2 | 4 | 0 | 20.00% | N | N |
| H2-30 | Colon | 10 | 0 | 9 | 0 | 90.00% | N | Y |
| H2-31 | Colon | 11 | 0 | 3 | 0 | 27.27% | N | Y |
| H2-32 | Colon | 3 | 0 | 12 | 0 | 400.00% | N | Y |
| H2-33 | Colon | 3 | 0 | 4 | 0 | 133.33% | N | N |
| H2-34 | Colon | 14 | 6 | 5 | 0 | 35.71% | N | N |
| H2-35 | Colon | 35 | 0 | 13 | 0 | 37.14% | N | N |
| H2-36 | Colon | 16 | 0 | 6 | 0 | 37.50% | N | N |
| H2-37 | Rectum | 5 | 0 | 2 | 0 | 40.00% | N | N |
| H2-38 | Colon | 11 | 3 | 0 | 0 | 0.00% | N | N |
| H2-39 | Stomach | 11 | 0 | 0 | 0 | 0.00% | N | N |
| H2-40 | Rectum | 20 | 2 | 4 | 0 | 20.00% | N | N |
| H2-41 | Colon | 9 | 0 | 3 | 0 | 33.33% | N | Y |
| H2-42 | Rectum | 10 | 2 | 6 | 0 | 60.00% | N | Y |
| H2-43 | Colon | 10 | 0 | 2 | 0 | 20.00% | N | Y |
| H2-44 | Colon | 2 | 0 | 1 | 1 | 50.00% | Y | N |
| H2-45 | Colon | 4 | 0 | 1 | 0 | 25.00% | N | N |
| H2-46 | Colon | 5 | 2 | 3 | 0 | 60.00% | N | N |
| H2-47 | Colon | 4 | 0 | 2 | 0 | 50.00% | N | N |
| H2-48 | Colon | 10 | 0 | 2 | 0 | 20.00% | N | Y |
| H2-49 | Rectum | 9 | 2 | 0 | 0 | 0.00% | N | N |
| H2-50 | Colon | 1 | 0 | 3 | 0 | 300.00% | N | N |
| H2-51 | Colon | 6 | 0 | 5 | 0 | 83.33% | N | N |
| H2-52 | Rectum | 8 | 2 | 4 | 2 | 50.00% | Y | Y |
| H2-53 | Colon | 6 | 6 | 0 | 0 | 0.00% | N | N |
| H2-54 | Colon | 1 | 0 | 3 | 0 | 300.00% | N | N |
| H2-55 | Colon | 12 | 7 | 0 | 0 | 0.00% | N | N |
| H2-56 | Colon | 3 | 0 | 1 | 0 | 33.33% | N | N |
| H2-57 | Stomach | 5 | 0 | 3 | 0 | 60.00% | N | N |
| H2-58 | Colon | 0 | 0 | 2 | 0 | 100.00% | N | N |
| H2-59 | Colon | 13 | 0 | 5 | 0 | 38.46% | N | N |
| H2-60 | Colon | 2 | 0 | 3 | 0 | 150.00% | N | N |
| H2-61 | Colon | 18 | 0 | 3 | 0 | 16.67% | N | N |
| H2-62 | Colon | 9 | 0 | 0 | 0 | 0.00% | N | N |
| H2-63 | Colon | 5 | 0 | 2 | 0 | 40.00% | N | N |
| H2-64 | Colon | 25 | 0 | 3 | 0 | 12.00% | N | N |
| H2-65 | Rectum | 15 | 4 | 0 | 0 | 0.00% | N | N |
| H2-66 | Colon | 3 | 0 | 3 | 0 | 100.00% | N | N |
| H2-67 | Colon | 12 | 0 | 2 | 0 | 16.67% | N | N |

### Improvement in lymph node counts

As shown in Table 3, gross examination assisted by shortwave infrared imaging identified a significant number (nonzero) of lymph nodes in residual tissue after standard of care lymph node dissection. b = 1.57, *X* ^2^= 1232.79, p < 0.0001.

**Table 3.** Comparison of nodal yield with traditional palpation and with shortwave infrared imaging.

|  | Traditional Palpation | Shortwave infrared-assisted Grossing |
| --- | --- | --- |
| **N Cases Reviewed with non-missing values** | 104 | 104 |
| **Total Nodes Identified** | 1165 | 500** |
| **Mean Nodes Identified (SD)** | 11.20 (10.74) | 4.81 (4.54) |
| **Mean Percent Improvement (% calculated for each case)\*** |  | 79.6% |
| **Total Positive Nodes** | 104 | 14*** |
| **Mean Positive Nodes (SD)** | 1.0 (2.57) | 0.13 (0.42) |

\* Mean case total % Improvement = mean of (lymph nodes count identified in Batch B per case/ lymph node count identified in Batch A per case).
\*\* Shortwave infrared imaging helped identify a significant number of nodes after traditional grossing, $b = 1.57$ , $= 1232.79$ , $p < 0.0001$ .
\*\*\* Shortwave infrared imaging helped a significant number of positive nodes after traditional grossing, $b = -2.01$ , $= 56.30$ , $p < 0.0001$

As shown in Table 4, in 36 of the 37 cases (97.3%) in Hospital 1 and 54 of the 67 cases (80.6%) in Hospital 2, additional lymph nodes were identified with the assistance of shortwave infrared imaging. Overall, in 90 of the 104 cases (86.5%), additional lymph nodes were identified with the assistance of shortwave infrared imaging.

**Table 4.** Comparison of hospital 1 and hospital 2.

|  | Hospital 1 | Hospital 2 | Total |
| --- | --- | --- | --- |
| **Total number of cases** | 37 | 67 | 104 |
| **Number of cases where shortwave infrared imaging helped identify additional lymph nodes** | 36 | 54 | 90 |
| **Percentage of cases where shortwave infrared imaging helped identify additional lymph nodes** | 97.3% | 80.6% | 86.5% |
| Number of cases overall positive cases | 8 | 23 | 31 |
| Numbers of cases where shortwave infrared imaging helped identified additional positive lymph nodes | 4 | 7 | 11 |
| Percentage of cases where shortwave infrared imaging identified additional positive lymph nodes | 10.8% | 10.4% | 10.6% |
| Percentage of positive cases where shortwave infrared imaging identified additional positive lymph nodes | 50.0% | 30.4% | 35.5% |
| Number of colorectal cases | 25 | 60 | 85 |
| Number of colorectal cases where LN compliance was not met after the first round* | 13 | 43 | 56 |
| Number of colorectal cases where LN compliance was met after shortwave infrared imaging-assisted gross examination | 7 | 13 | 20 |
| Percentage of cases where LN compliance was improved due to shortwave infrared imaging assisted gross examination | 53.9% | 30.2% | 35.7% |

\*LN means Lymph Node
\*\*LN compliance means the requirement of a minimum of twelve lymph nodes examined for colorectal cancer cases.

### Improvement in positive lymph node counts

As shown in Table 3, gross examination assisted by shortwave infrared imaging identified a significant number (nonzero) of positive lymph nodes in samples discarded after standard of care lymph node dissection, b = -2.01, *X* ^2^= 56.30, p < 0.0001.

As shown in Table 4, in 4 of the 37 cases (10.8%) in Hospital 1 and 7 of the 67 cases (10.4%) in Hospital 2, additional positive lymph nodes were identified with the assistance of shortwave infrared imaging. Overall, in 11 of the 104 cases (10.6%), additional lymph nodes were identified with the assistance of shortwave infrared imaging.

There were 31 lymph node positive cases in total (8 from Hospital 1 and 23 from Hospital 2). In 4 of these positive cases (12.9%), the positive lymph nodes were only discovered by shortwave infrared imaging, with no positive lymph nodes being identified through the standard of care visual and manual dissection. These 4 positive cases would have been wrongly staged as lymph node negative in the absence of shortwave infrared imaging support. An example where the only positive lymph node was identified with the help of shortwave infrared imaging was depicted in Figure 1(B) and (C).

Also, in 11 of the 31 the lymph node positive cases (35.5%), additional lymph nodes were identified with the assistance of shortwave infrared imaging.

## DISCUSSION

The main purpose of this study was to evaluate the effectiveness of shortwave infrared imaging in (1) increasing lymph node counts and (2) identifying additional positive lymph nodes. As shown in the **RESULTS** section, both hypotheses were strongly supported.

In addition to supporting these two main hypotheses, we also investigated the effectiveness of shortwave infrared imaging for improving compliance of gross examination of colorectal cancer cases. Established guidelines in the United States dictate that colorectal cancer cases yield a minimum of twelve lymph nodes. Failure in reaching twelve lymph nodes after the first round of gross examination results in re-examination of residual adipose tissue, which is time-consuming, labor intensive, and economically costly.(15) This problem is particularly common for cases that had been previously treated with radiation or chemotherapy.(2) Lymph nodes often become smaller and are thus more challenging to retrieve after such treatments, with reassessment of the gross specimen multiple times being common for such cases. In this study, only 34.1% (29/85) of colorectal cancer cases achieved the required number of 12 lymph nodes after the first round of grossing. Shortwave infrared imaging brought 35.7% (20/56) of the non-compliant cases into compliance, obviating the need to return to the specimen in over a third of these cases.

The data provides compelling evidence for the benefit of shortwave infrared imaging in lymph node dissection in the surgical pathology laboratory. However, as a first large scale multi-center pilot study of this emerging technology, there are several limitations to this study.

The first limitation is the inconsistency in the user proficiency in the technology. When comparing the percentage improvement that resulted from the assistance of shortwave infrared imaging in (1) increasing lymph node counts, (2) increasing positive lymph node counts, and (3) improving compliance, Hospital 1 consistently showed a larger improvement, which could suggest a higher proficiency level of the users in Hospital 1. Because Cision InVision™ Pathology Imaging System does not directly identify lymph nodes (but rather shows a shortwave infrared reflection which is indicative of the natural water content difference between tissue types), the users are responsible for interpreting the images in real time and deciding on which tissue pieces to submit. Robust training and testing protocols for users in a future study can help overcome this limitation.

The second limitation is that the two-step evaluation process is not representative of the practical workflow for using the shortwave infrared imaging system. In a real-world scenario, the first round of gross examination can be carried out together with the assistance of the shortwave infrared imaging system. There is no practical reason for the user to only use the device for searching additional lymph nodes in residual adipose tissue after manual palpation is completed. One can argue that the shortwave infrared imaging system can have an even larger impact in a real-world scenario compared to what has been shown in this study, because the two-step approach places this new technology in a disadvantaged position to demonstrate its effectiveness. Future studies where shortwave infrared imaging is used in the first round of gross examination may be conducted to measure the effectiveness of this new technology in standard practice settings.

The third limitation is that this study was not completely blinded. Although we blinded the pathologists while they review histological results from different batches, there is no way to blind the prosectors with regard to gross examination with or without the device. The psychological factors of the users of the device could have played a role in affecting the numbers of lymph node counts. However, because these procedures were conducted all by experienced board-certified PAs with patients’ best interests in mind, the impact of this role is unlikely to have changed any of the major conclusions that we drew from the data above.

Other potential effects that shortwave infrared imaging can have that were not investigated in this study are (1) time saving in lymph node searches and (2) cost saving for reducing the number of cassettes submitted for histological processing. Future studies need to be conducted to evaluate these potential benefits.

In conclusion, this two-center, 104-patient large-scale pilot study, demonstrated that shortwave infrared imaging helps not only find additional lymph nodes, but also additional positive lymph nodes. Future studies will need to be conducted to overcome the limitations of this study and to investigate other potential benefits of shortwave infrared imaging.

## Data Availability

All data produced in the present work are contained in the manuscript.

## Competing Interests

S Khelifa, R Seltzer, J Manuel, M W Rosenbaum, J Almas, Z Li are shareholders and/or employees of Cision Vision, Inc., a company that sells shortwave infrared imaging systems, including Cision InVision™ Pathology Imaging System.

## Notes

### Funding Statement

This study did not receive any funding

### Author Declarations

IRB of Wisconsin Medical College gave ethical approval for this work. IRB of Franciscan Missionaries of Our Lady Health System gave ethical approval for this work.

